# Impact of the COVID-19 Pandemic and Vaccine Hesitancy among Farmworkers from Monterey County, California

**DOI:** 10.1101/2020.12.18.20248518

**Authors:** Ana M. Mora, Joseph A. Lewnard, Katherine Kogut, Stephen Rauch, Norma Morga, Nicholas Jewell, Maximiliano Cuevas, Brenda Eskenazi, on behalf of the CHAMACOS/Project-19 Study Team

## Abstract

**Objectives:** To examine the impact of the COVID-19 pandemic on farmworkers from Monterey County, California.

**Methods:** We recruited adult farmworkers (n=1115) between July 16, 2020 and November 30, 2020. We collected information on sociodemographic characteristics, health behaviors, economic and social stressors experienced during COVID-19, and willingness to be vaccinated via interviews by phone.

**Results:** Study participants, particularly female farmworkers, reported adverse effects of the pandemic on their mental health and home environment (e.g., 24% overall reported depression and/or anxiety symptoms). The pandemic also resulted in greater financial burden for many farmworkers, with 37% food insecure and 51% unable to pay bills. Half of respondents reported that they were extremely likely to be vaccinated. Vaccine hesitancy was most common in participants who were women, younger, born in the United States, and living in more rural areas.

**Conclusions:** We found that the pandemic has substantially impacted the mental and physical health and economic and food security of farmworkers.

**Public Health Implications:** This study highlights the need to provide farmworkers with supplemental income, and increased mental and family health, and food support services.

## INTRODUCTION

Latinos have accounted for a disproportionate share of COVID-19 cases in the United States with 5-7 times higher COVID-19-related mortality than non-Hispanic whites.^1^ United States farmworkers, most of whom are Latino, have been considered essential workers during the COVID-19 pandemic due to their role ensuring continuity of the country’s food supply. California is the leading state in terms of agricultural production and employs approximately 800,000 farmworkers,^2^ who largely reside in the central and coastal valleys, including Fresno, Kern, and Monterey counties.^3,4^ About 90% of California farmworkers are born in Mexico,^5^ at least 50% are undocumented,^7^ approximately 20% speak indigenous languages,^6^ an estimated half are uninsured,^7^ and most live within 200% of federal poverty level.^6^

In Monterey County, California (as of November 30, 2020), 74% of COVID-19 cases were Latinos — a prevalence in excess of its 59% Latino population; 19% of these cases were farmworkers.^8,9^ Apart from increased infection, concerns have arisen that the living and working conditions experienced by farmworkers may exacerbate the psychological, economic, and social impact of the pandemic on farmworkers. Herein, we describe this impact as reported by 1115 farmworkers in Monterey County in a cross-sectional study. We determined which subgroups of farmworkers were most impacted, and whether different demographic and social factors predict their willingness to be vaccinated.

## METHODS

This study was undertaken through a collaboration between Clinica de Salud del Valle de Salinas (CSVS), a federally qualified health center, and University of California at Berkeley School of Public Health. Recruitment for this study occurred between July 16, 2020 and November 30, 2020. Individuals were eligible to participate if they were age 18 years or older, spoke Spanish or English, were not pregnant, were receiving a test for SARS-CoV-2 infection at CSVS, and had not tested positive for SARS-CoV-2 in the past two weeks. From July 16 to October 7, we limited enrollment to individuals who had engaged in agricultural work in the past two weeks. Due to the ending of the harvest season, from October 8 onward we enrolled anyone who had worked in agriculture since March 2020. Recruitment took place on-site at CSVS clinics (n=565) or at community outreach events (n=550), including community health fairs and visits to farmworker housing complexes. A total of 1115 farmworkers were enrolled in the study.

Interviews were conducted by phone by bilingual and bicultural research assistants drawn from the local Salinas Valley community. Interviews, using a structured questionnaire, were usually completed on the same day as SARS-CoV-2 testing (N=739) or the following day (N=292); only a few respondents knew their testing result at the time of interview. Surveys were conducted in Spanish (N=1000) or English (N=115) and covered sociodemographic characteristics; employment information; risk factors for SARS-CoV-2 infection and safety practices at home, in the community, and in the workplace; COVID-19 symptoms; other medical conditions and health behaviors, including smoking and substance use; and economic and social stressors experienced during the COVID-19 pandemic.

Protocols were approved by the Office for the Protection of Human Subjects at UC Berkeley. All participants provided informed written consent after having the opportunity to hear the consent presented aloud in either Spanish or English. Participants received a $50 cash-value debit card as their incentive for participating in the study.

## Data analysis

We considered the following demographic and household characteristics (Table 1): sex (male vs female), age (continuous), language spoken (Spanish, English, or indigenous languages), country of birth (Mexico, United States, or other), years spent in the United States prior to the study (for non-native residents; continuous), place of residence (Salinas, Greenfield, or other), education level (completed only primary school or less vs. more than primary), annual household income (<$25,000 vs. ≥$25,000), marital status (married or living with a partner vs not married or living with a partner), overcrowded housing (as defined by the United States Department of Housing and Urban Development; ≤2 persons per bedroom vs >2 persons per bedroom),^10^ living with unrelated roommates (yes vs no), living with children under 18 (yes vs no), type of agricultural work performed (field work vs other), and recruitment site (clinic vs community outreach event). Participants who were bilingual in Spanish and an indigenous language were considered to be indigenous speakers; bilingual in Spanish and English were considered to be Spanish speakers. We examined the prevalence of these sociodemographic characteristics in the full study population, and additionally assessed differences by sex or recruitment site (clinic vs outreach) using chi-square tests (for categorical variables) or t-tests (for continuous variables).

**Table 1.**
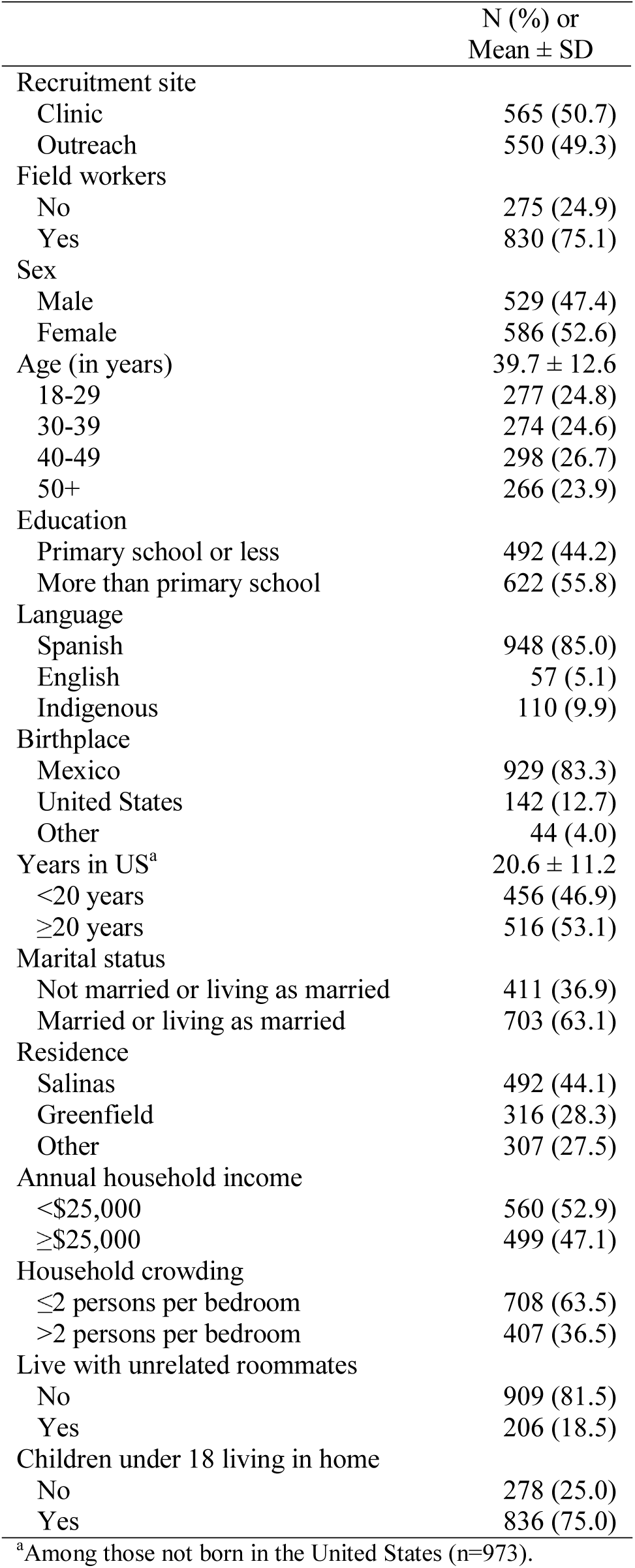
Demographic and household characteristics, Monterey County COVID-19 Farmworker Study, July to November 2020, N=1115.

We aimed to assess multiple aspects of participants’ well-being in the context of the pandemic. To ascertain symptoms of depression, we used the Patient Health Questionnaire-2 (PHQ-2)^11^ and for symptoms of anxiety, we used the Generalized Anxiety Disorder-2 (GAD-2) scale.^12^ We defined symptoms of depression as a score of ≥2 on the PHQ-2^13^ and symptoms of anxiety as a score of ≥2 on the GAD-2.^14^ We also created a composite measure of depression and/or anxiety symptoms. We asked a number of Yes/No questions about participants’ feelings and behaviors “compared to before the COVID-19 pandemic.” These included whether they “felt very fearful of catching COVID-19 (for yourself or others)”, “felt unhappy with your life”, “had difficulty sleeping”, and many other questions (see Table 2). We asked whether they had increased their use of alcohol, tobacco, marijuana, and other substances like pills or other drugs; we also created a composite measure indicating an increase in use of any of these substances. For assessment of food insecurity, we used a slight modification of the six-question scale developed by the United States Department of Agriculture (USDA),^15^ altering the time period to be “since the pandemic started” rather than the last 12 months. Levels of food security were defined using USDA cut-offs for high, marginal, low, or very low food security; for the purposes of analysis, we collapsed the two lowest food security groups and compared to the two highest. In addition, we added a global question “do you feel that your ability to buy food for you and/or your household is the same as, less than, or more than before the COVID-19 pandemic?” and we recoded responses for analyses as “Yes, less” or “No”. Midway through the study we added the question “Have you had more difficulty paying your bills (including water, gas and electricity, rent, and childcare) since the COVID-19 pandemic started?”; hence the sample size is smaller for this question. We asked whether participants had sent remittances to family members outside the United States just prior to the pandemic, and amongst those who had, we asked whether they were now sending less, more, or the same; responses were recoded as “Yes, less” or “No”. Lastly, to better understand the participants’ own assessment of impacts of the pandemic, we asked general questions: “How much of a negative impact the COVID-19 has had on your life?” and “How concerned are you about COVID-19?” COVID-19 impacts were considered for the full study population as well as stratified by either sex or recruitment site (clinic vs outreach); differences between strata were assessed using chi-square tests.

**Table 2.**
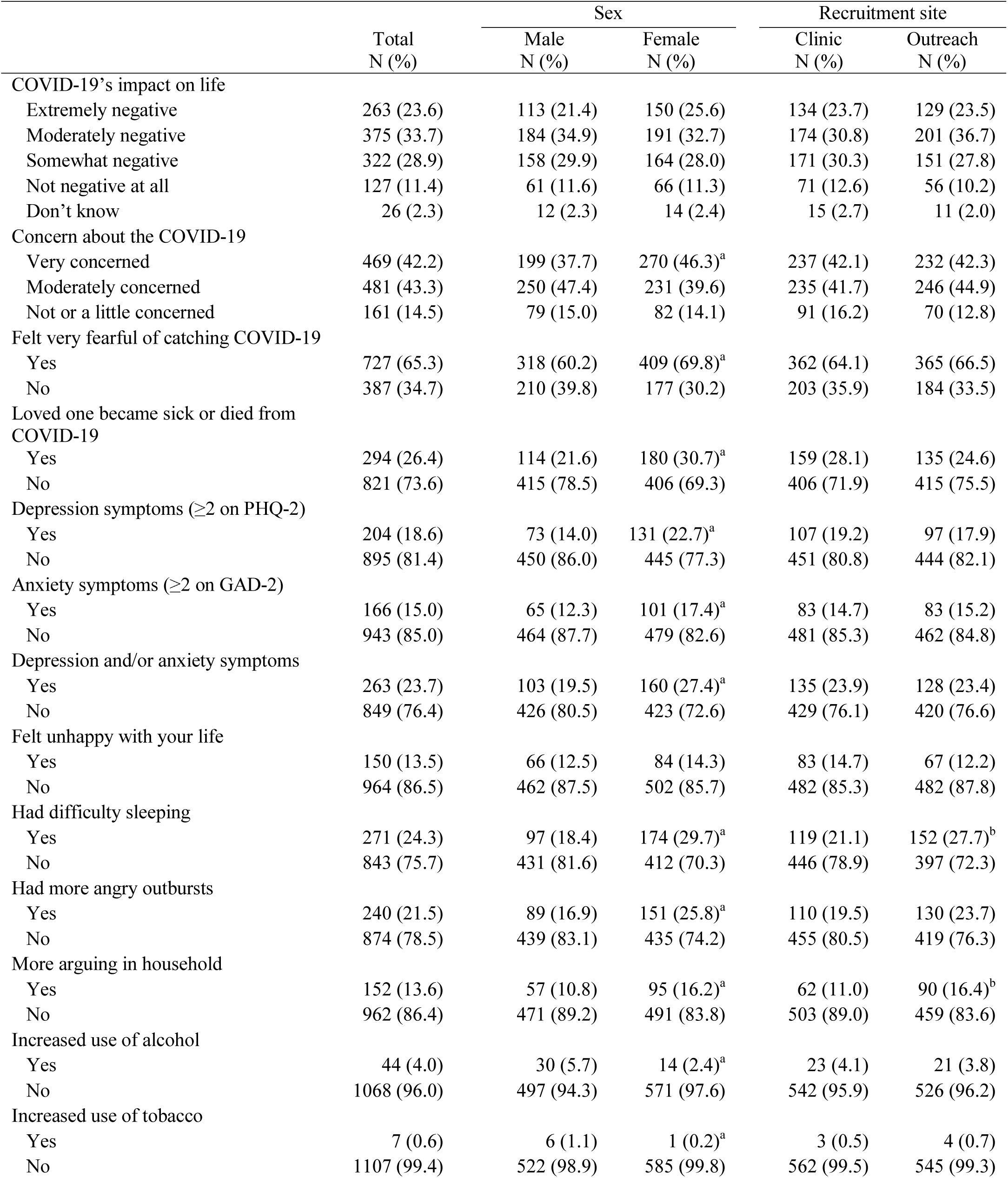
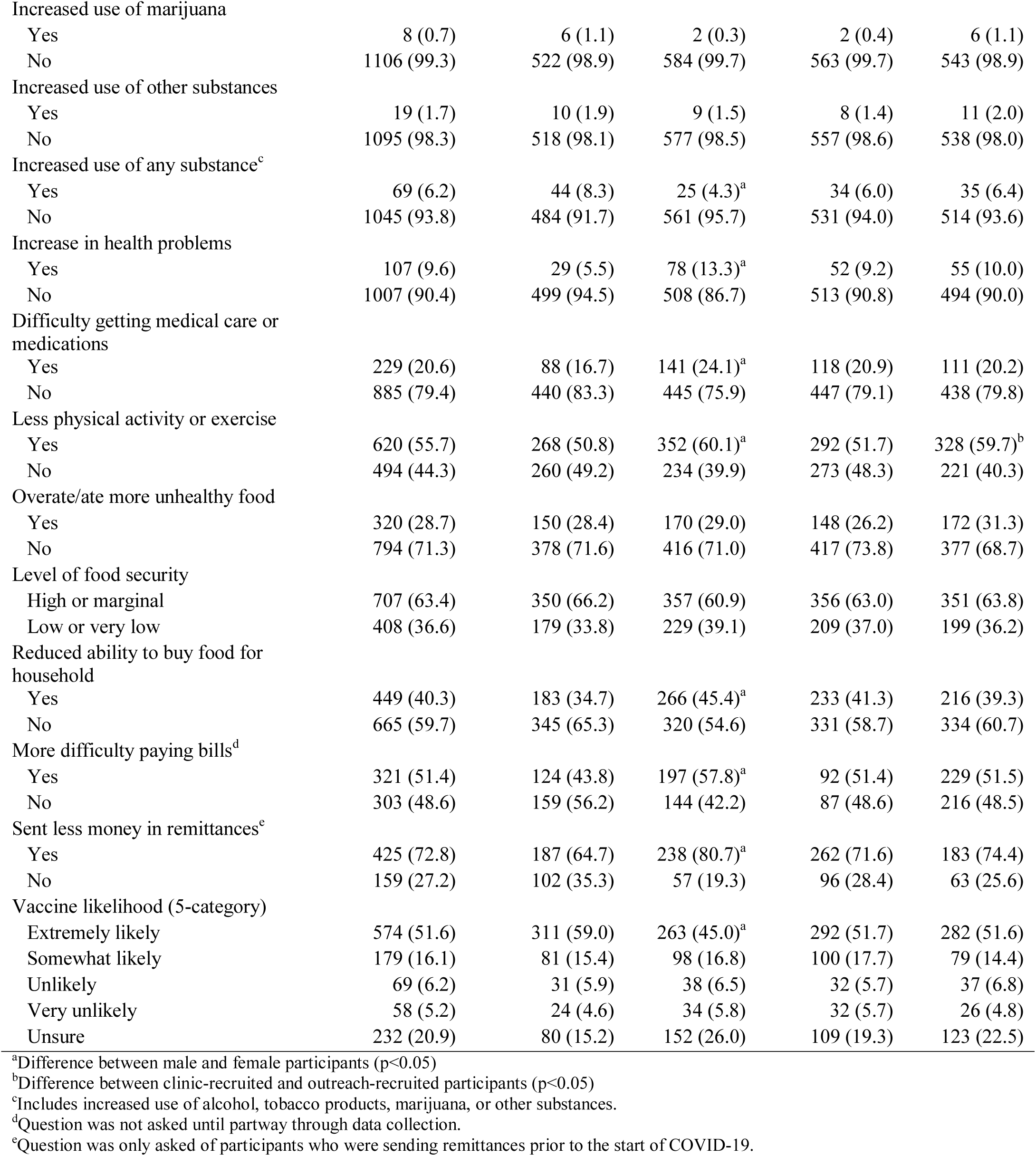
Impacts of COVID-19 pandemic on participants in Monterey County COVID-19 Farmworker Study, July to November 2020, N=1115.

In addition, we assessed participants’ intentions to get the COVID-19 vaccine: “How likely are you to get vaccinated for COVID-19 once a vaccine is available to you?” Participants who did not indicate that they were extremely likely to be vaccinated were asked reasons for why they would not get vaccinated. We performed bivariate analyses to examine the relationship of the demographic and household characteristics, as listed above, with the additional inclusion of whether they had a loved one who became sick or died from COVID-19, as possible predictors of intention to get vaccinated. We used chi-square tests (for categorical predictors) or t-tests (for continuous predictors) to explore associations. To assess independent predictors of participants’ willingness to be vaccinated, we fit a relative risk model using Stata’s GLM command with Poisson family and log link,^16^ with the outcome of a participant reporting not being extremely likely to be vaccinated, and including all covariates considered in bivariate analyses. We used Huber-White robust standard errors to account for model misspecification. Current age and years spent in the United States were entered as continuous variables and were scaled to identify the change in risk associated with a 10-year duration of stay in the United States. Participants who were born in the United States were assigned their current age as a measure of the time spent in the country.

## RESULTS

Table 1 describes the sociodemographic characteristics of the study participants. Most worked in the fields (75%) and resided in Salinas (44%) or Greenfield (28%) in Monterey County. The average age of respondents was 39.7 (Standard Deviation, SD=12.6) years, 47% were male, and 63% were married or living with a partner. Education level was low with nearly half (44%) having only primary school or lower levels of attainment. Most participants were from Mexico. Among respondents who were born outside of the United States, the average length of time in the United States was 20.6 (SD=11.2) years. Most farmworkers (85%) spoke Spanish at home, although 10% reported speaking indigenous languages such as Mixteco, Triqui, or Zapoteco. About half (53%) of the respondents reported earning less than $25,000 per year. Many respondents (37%) lived in overcrowded housing and three-quarter of the homes had children younger than 18 years old.

Many farmworkers (42%) reported that they were very concerned about the pandemic, with 65% reporting feeling fearful of falling ill with COVID-19 (Table 2). About 26% reported that a loved one became sick or died from COVID-19. Nearly a quarter reported symptoms of depression and/or anxiety, having difficulty sleeping, or having more angry outbursts. When asked in comparison to before the pandemic, 21% reported having more difficulty getting needed medical care or medications, 56% reported less physical activity, 29% reported overeating or eating less healthy foods, and 6% reported increasing their use of cigarettes, marijuana, alcohol, or other substances like pills or drugs. A large proportion of the farmworkers reported that, since the pandemic started, they had experienced food insecurity (37%), including 9% who met the threshold of very low food security. In comparison to before the pandemic, 40% reported that they were less able to buy food for themselves or their household. In general, half of the farmworkers reported having more difficulty paying bills since the pandemic started and the economic toll was also reflected in lower contributions to family members abroad. Specifically, among the 54% of farmworkers who had been sending remittances to support family members outside the United States prior to the pandemic, 73% now sent less. Overall, 24% reported they were extremely negatively impacted by COVID-19.

We observed differences in the reporting of adverse impacts by men and women (Table 2). As compared to men, a higher proportion of women reported being very concerned about the pandemic (46% women vs. 38% men) and about catching COVID-19 (70% women vs. 60% men). Women were also more likely than men to report symptoms of depression and/or anxiety, difficulty sleeping, more angry outbursts and more arguing in the household, having a loved one sick with COVID-19, an increase in health problems, difficulty getting medical care or medications, less physical exercise, reduced ability to buy food, more difficulty paying bills, and sending less money in remittances. Although numbers are small, men were more likely than women to report an increase in their use of alcohol or tobacco. There were a few differences between those who were recruited from the clinics and those from outreach. Specifically, those recruited from outreach were more likely to report having more difficulty sleeping, having more arguing in the household, and getting less physical activity or exercise compared to those recruited from clinics.

Only 52% of respondents reported that they were extremely likely to be vaccinated once a vaccine was available to them and 32% were unsure or unlikely to be vaccinated (Table 3). The main reason participants reported for not being extremely likely to be vaccinated (Figure 1) was concern about side effects or getting COVID-19 from the vaccination; other less common yet important deterrents to vaccination were distrust of the government, not understanding what a vaccine was, or not believing in its utility. As shown in Table 3, some subgroups of farmworkers had lower enthusiasm towards vaccination than others. For example, females and those who were younger, born in the United States, and living in the more rural towns in south Monterey County were less likely to wish to be vaccinated.

**Table 3.** Sociodemographic factors associated with likelihood of getting vaccinated, Monterey County COVID-19 Farmworker Study, July to November 2020, N=1115.

| | Extremely likely to get vaccinated<br>N (%) or Mean ± SD | Not extremely likely to get vaccinated<br>N (%) or Mean ± SD | RR (95% CI) of not extremely likely to get vaccinated<sup>d</sup> |
| --- | --- | --- | --- |
| All participants | 574 (51.6) | 538 (48.4) | N/A |
| Field workers |  |  |  |
| No | 145 (52.9) | 129 (47.1) | Ref |
| Yes | 422 (51.0) | 406 (49.0) | 1.02 (0.87, 1.18) |
| Sex |  |  |  |
| Male | 311 (59.0) | 216 (41.0)<sup>a</sup> | Ref |
| Female | 263 (45.0) | 322 (55.0) | 1.37 (1.20, 1.57)<sup>a</sup> |
| Age in years | 41.7 ± 12.7 | 37.6 ± 12.1<sup>a</sup> | 0.86 (0.79, 0.95)<sup>a,b</sup> |
| 18-29 | 118 (42.9) | 157 (57.1) |  |
| 30-39 | 129 (47.1) | 145 (52.9) |  |
| 40-49 | 163 (54.7) | 135 (45.3) |  |
| 50+ | 164 (61.9) | 101 (38.1) |  |
| Education |  |  |  |
| Primary school or less | 266 (54.2) | 225 (45.8) | Ref |
| More than primary school | 308 (49.7) | 312 (50.3) | 0.98 (0.84, 1.14) |
| Language |  |  |  |
| Spanish | 500 (52.9) | 446 (47.2) | Ref |
| English | 23 (40.4) | 34 (59.7) | 1.00 (0.78, 1.30) |
| Indigenous | 51 (46.8) | 58 (53.2) | 1.04 (0.82, 1.31) |
| Birthplace |  |  |  |
| Mexico | 497 (53.6) | 430 (46.4)<sup>a</sup> | Ref |
| United States | 50 (35.5) | 91 (64.5) | 1.12 (0.89, 1.41) |
| Other | 27 (61.4) | 17 (38.6) | 0.83 (0.57, 1.21) |
| Years in US | 21.4 ± 11.8 | 19.8 ± 10.5<sup>a,c</sup> | 1.03 (0.93, 1.15)<sup>b</sup> |
| <20 years | 222 (48.8) | 233 (51.2) |  |
| ≥20 years | 302 (58.6) | 213 (41.4) | |
| Marital status |  |  |  |
| Not married or living as married | 192 (46.9) | 217 (53.1)<sup>a</sup> | Ref |
| Married or living as married | 381 (54.3) | 321 (45.7) | 0.94 (0.82, 1.08) |
| Residence |  |  |  |
| Salinas | 268 (54.6) | 223 (45.4)<sup>a</sup> | Ref |
| Greenfield | 145 (45.9) | 171 (54.1) | 1.19 (1.02, 1.39)<sup>a</sup> |
| Other | 161 (52.8) | 144 (47.2) | 1.04 (0.89, 1.22) |
| Annual household income |  |  |  |
| <\$25,000 | 281 (50.5) | 276 (49.6) | Ref |
| ≥\$25,000 | 268 (53.7) | 231 (46.3) | 1.05 (0.91, 1.20) |
| Household crowding |  |  |  |
| ≤2 persons per bedroom | 368 (52.1) | 339 (48.0) | Ref |
| >2 persons per bedroom | 206 (50.9) | 199 (49.1) | 0.99 (0.87, 1.14) |
| Live with unrelated roommates |  |  |  |
| No | 463 (51.1) | 444 (49.0) | Ref |
| Yes | 111 (54.2) | 94 (45.9) | 0.95 (0.79, 1.14) |
| Children under 18 living in home |  |  |  |
| No | 158 (57.0) | 119 (43.0)<sup>a</sup> | Ref |
| Yes | 416 (49.9) | 418 (50.1) | 1.02 (0.86, 1.20) |

<sup>a</sup>Different at $p < 0.05$
<sup>b</sup>Relative risk per 10-year difference
<sup>c</sup>Among those not born in the United States (n=973).
<sup>d</sup>n for the multivariate model=1045 due to 54 “Don’t know” responses to the income question and missing values for field worker status (n=10), education (n=1), marital status (n=1), children living at home (n=1), and vaccine outcome (n=3).

**Figure 1.**
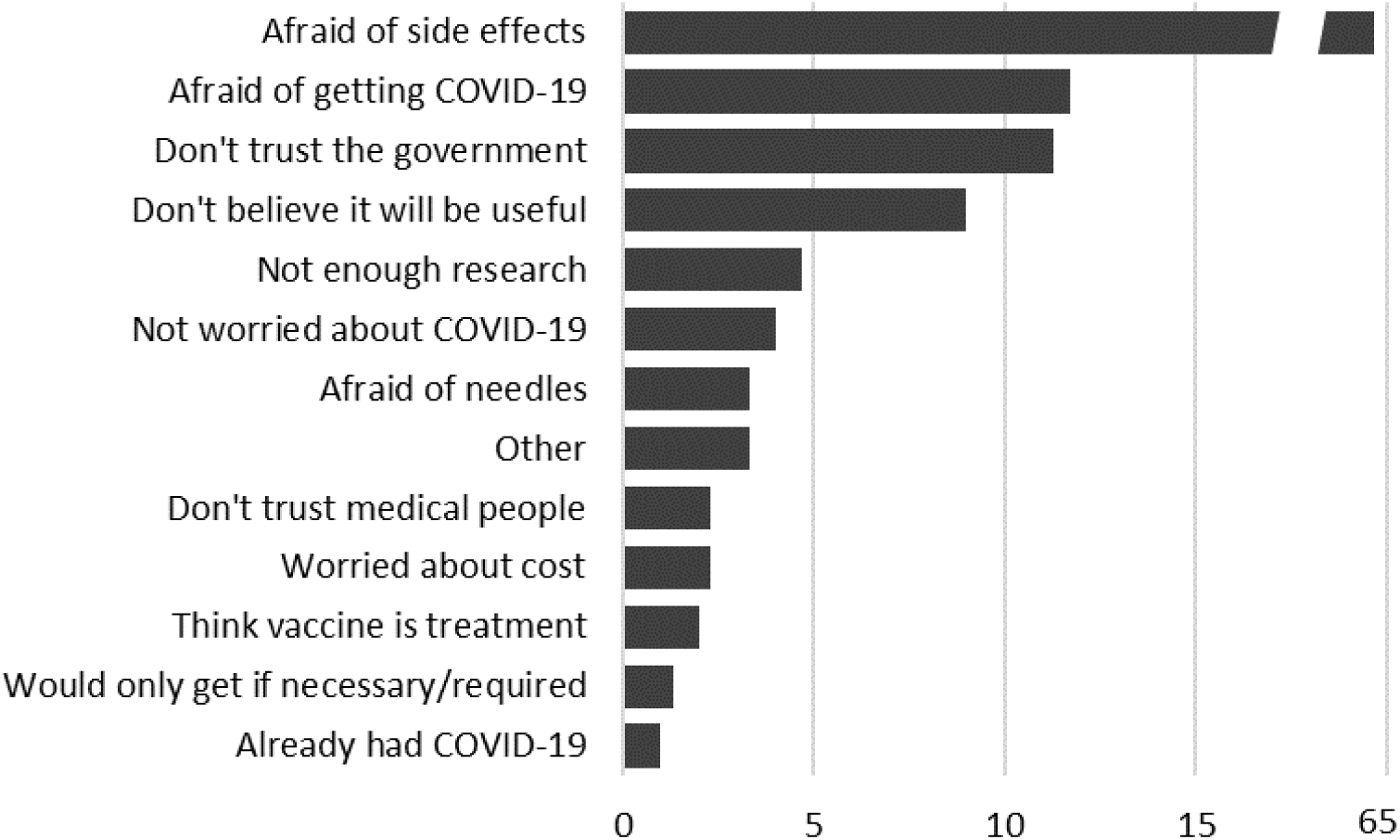
Reasons provided by farmworkers for why they were not extremely likely to get vaccinated, Monterey County COVID-19 Farmworker Study, July to November 2020, N=300.

## DISCUSSION

Our study of farmworkers in Monterey County, California reveals impacts of the COVID-19 pandemic extending beyond infection. We observed that a significant number of our study participants reported adverse effects of the pandemic on their mental health and home environment, particularly female farmworkers. Additionally, farmworkers reported that the pandemic may have potentially affected their physical health by preventing some workers from receiving medical care or medications, getting physical exercise, and eating right. While numbers were low, some farmworkers, particularly men, reported increased substance use. The pandemic has also resulted in greater financial burden for many farmworkers, as indicated by their reduced ability to buy food, pay bills, and send money to family abroad. The 37% prevalence of food insecurity reported by farmworkers in our study is more than three times greater than the pre-pandemic estimate of 10.5% in the United States population at large,^17^ but is comparable to national estimates of food insecurity early in the pandemic.^18,19^ The stay-at-home directives to limit socializing with friends and family, and concerns about whether they or loved ones will catch COVID-19 likely contribute to the observed reports of mental health problems or more hostile home environments. Both the stay-at-home restrictions and high concern about getting COVID-19 have likely led to reduced physical activity or access to medical care.

Our findings suggest that impacts of the COVID-19 pandemic on the economic losses of the agricultural sector trickled down to the farmworkers. By May 2020, California farmers lost an estimated $2 billion due to the pandemic, with an additional estimated loss of $6 billion projected through the end of the year,^20^ emanating from disruptions in export markets, distribution supply chains, reductions in the food service industry, and consumers’ shift to shelf-stable items.^20^ In Monterey County alone, farmworker employment decreased 40% in the period between April 2019 and 2020,^20^ with similar job losses in agricultural counties throughout the state.^20^ This was reflected in the greater economic hardships reported by farmworkers in this study. More than half of the Monterey County farmworkers we surveyed reported an increase in inability to pay bills and 40% reported reduced ability to buy food. This proportion was even greater in the 915 farmworkers from the statewide COVID-19 Farmworkers’ Study (COFS),^21^ who were surveyed earlier in the pandemic (May and July 2020). More specifically, in COFS, 63% of farmworkers had more difficulty paying rent, 60% paying utilities, and 70% buying food during the pandemic (Rick Mines, Personal Communication, December 5, 2020), and overall, 52% of farmworkers reported a decrease in employment.^21^ The economic impact on farmworkers and their families is not limited to California in that smaller studies in Oregon^22^ and North Carolina^23^ reported similar loss of employment and economic hardship among farmworkers.

Although little is known about the differential impacts of epidemics on men and women, data from the Ebola outbreak in West Africa and the Zika outbreak in Brazil indicate that women bear the greatest social, emotional, and economic burden during infectious-disease outbreaks.^24^ For example, women carry most care responsibilities when family members fall ill, are at greater risk of domestic violence, and are also more affected by job losses in times of economic instability.^25–27^ In our study of farmworkers, we observed that women were more likely than men to report fears and concerns about COVID-19, mental health symptoms, and financial difficulties. This could be due to the fact that, compared to men, women are more likely to *report* the health effects such as depression and anxiety^28^ in addition to the possibility that they are more likely to *experience* these impacts, as described above.

A limitation of the current study is that we were not able to ask farmworkers about all the potential effects of this pandemic. For example, we did not ask about what preparations were made to care for minor children and what pressure this imposed on the families. We know that 75% of the households had children under 18 years and 37% had children 5 years old or younger (some may not have been the respondent’s children). Very few schools and childcare facilities were open during the pandemic and only 8% of the children were in school or childcare. During the pandemic, parents had the additional responsibility for facilitating the education of their school-age children, for which many of the farmworkers in our study may not have had the proper educational, technological, or language skills. Affordable childcare is difficult to find in the best of times for these farmworker families.^29^ Anecdotal stories from the Salinas Valley community suggest that women were more likely to leave their jobs to provide the needed childcare and educational support for their children, resulting in even greater economic hardship, especially for single parents, but also in greater emotional burden for women.

Another limitation of this study was that the results may not be generalizable to farmworkers across California, the United States, or in other countries. Our study was not a random sample of the farmworker population, and the National Agricultural Workers Survey (NAWS) of the Department of Labor, the only random-sample survey of California or United States crop workers, was last conducted in 2015-2016.^30^ Because of our concern about potential bias in a clinic-based population, we recruited about half of the participants from the community through health fairs, housing facilities for farmworkers, leaflets, and employers, and we found that those recruited from outside the clinic reported greater effects of the pandemic (i.e., difficulty sleeping, more arguing in the household, less physical exercise). Still, we may not have reached those most impacted by the pandemic. Fear of being reported to government authorities, of deportation,^31^ or of loss of other benefits (public charge) if found to be COVID-19 cases were likely to deter some farmworkers from being tested, especially among those who were undocumented, even though regulations were in place to prevent these punitive actions.^32,33^

As part of the essential workforce, farmworkers are likely to receive priority access to vaccination in California or other states. Our study has demonstrated the reticence of half of the farmworker population to be vaccinated to prevent COVID-19. In a Pew Research Center survey conducted November 18-29, 2020 of over 12,000 Americans, 60% said they would definitely or probably get vaccinated.^34^ In that study, Latinos were comparable to non-Hispanic Whites in their willingness to be vaccinated, but women and younger adults reported lower enthusiasm — findings consistent with ours. In our study, we found that those who lived in more rural communities and who were more acculturated (i.e., born in the United States) reported being less likely to get vaccinated. Although the main reason for not choosing to be vaccinated was concern about side effects, our results also show that distrust of the United States government as well as lower health literacy may play a role and need to be addressed in any vaccination program. The World Health Organization SAGE working group on vaccine hesitancy stated that overcoming hesitancy would require a multipronged approach, targeting specific populations that are most hesitant and addressing their specific concerns^35^ — as we have identified in the present study. In addition, a vaccination program for farmworkers should also consider the inherent difficulties in delivering vaccination to a “hidden” undocumented or uninsured population or to those who may be migratory.

In conclusion, we quantified the way in which the pandemic has substantially impacted the mental and physical health of farmworkers and their economic and food security. We have also identified that, in spite of this impact, a significant portion of the population remains reluctant to be vaccinated. Our findings suggest that interventions including providing supplemental income, increasing mental and family health services, and food support services may be of value for mitigating the impacts of the pandemic on this population. Because women may be experiencing the burden of the pandemic more acutely, we recommend services tailored to women in order to support them during this time. For those who test positive for SARS-CoV-2, we recommend immediate income replacement. Lastly, we recommend that vaccination programs for farmworkers are coupled with educational campaigns that target those with greatest resistance to vaccination and address the misunderstandings of vaccination and the distrust of the government. Utilizing respected and trusted members of the community, such as community-based organizations and clinics, will be essential for the success of these programs.

## Authors contributions

A.□ M. Mora, J. A. Lewnard, and B. Eskenazi led the conceptual development, writing, and editing of the manuscript. K. Kogut and N. Morga coordinated the study. S. Rauch conducted all data analyses. N. Jewell provided statistical consultation. M. Cuevas provided access to the population and significant clinical and logistical support in conducting the study. All authors provided critical feedback and edits on all aspects of the manuscript.

## Data Availability

Data are available on request.

## Acknowledgements

This work was supported by the Innovative Genomics Institute and Clinica de Salud del Valle de Salinas.

## Disclosures of potential and actual conflicts of interest

The authors declare that they have no competing interests.

## REFERENCES

1. Bassett MT, Chen JT, Krieger N. The unequal toll of COVID-19 mortality by age in the United States: quantifying racial/ethnic disparities. Harv Cent Popul Dev Stud Work Pap Ser. 2020;19(3).

2. Martin P, Hooker B, Akhtar M, Stockton M. How many workers are employed in California agriculture? Calif Agric. 2016;71(1):30–34.

3. Current Employment Statistics - CES.; 2020. https://www.bls.gov/ces/

4. California Department on Food & Agriculture. California Agricultural Production Statistics. https://www.cdfa.ca.gov/statistics/

5. Gabbard PS. Who Are California Crop Workers and How Is This Changing? Published online 2016. https://www.google.com/url?sa=t&rct=j&q=&esrc=s&source=web&cd=&cad=rja&uact=8&ved=2ahUKEwjjkYnRw9jrAhWRoFsKHb6cBGAQFjAJegQIAxAB&url=https%3A%2F%2Fdoleta.gov%2Fnaws%2Fresearch%2Fdocs%2FAbs_APMA_pres_Jan2016.pdf&usg=AOvVaw1bIrl8KKjqupv5gz0ujpBt (2016).

6. Mines R, Nichols S, Runsten D. California’s indigenous farmworkers. Final Rep Tof Indig Farmworker Study Califonria Endow. Published online 2010.

7. Hernandez T, Gabbard S. National Agricultural Workers Survey.; 2018. https://www.dol.gov/sites/dolgov/files/OASP/legacy/files/NAWS-Research-Report-13.pdf

8. Summary Report: 2019 Novel Coronavirus (2019-nCoV) - Local Data | Monterey County, CA. Published 2020. Accessed December 18, 2020. https://www.co.monterey.ca.us/government/departments-a-h/health/diseases/2019-novel-coronavirus-covid-19/2019-novel-coronavirus-2019-ncov-local-data-10219#sumaryreport

9. U.S. Census Bureau QuickFacts: Monterey County, California; Salinas city, California. Published 2019. Accessed December 18, 2020. https://www.census.gov/quickfacts/fact/table/montereycountycalifornia,salinascitycalifornia/ PST045219

10. Blake KS, Kellerson RL, Simic A. Measuring overcrowding in housing. Wash DC Dep Hous Urban Dev Off Policy Dev Res. Published online 2007.

11. Kroenke K, Spitzer RL, Williams JB. The Patient Health Questionnaire-2: validity of a two-item depression screener. Med Care. Published online 2003:1284-1292.

12. Kroenke K, Spitzer RL, Williams JB, Monahan PO, Löwe B. Anxiety disorders in primary care: prevalence, impairment, comorbidity, and detection. Ann Intern Med. 2007;146(5):317–325.

13. Arroll B, Goodyear-Smith F, Crengle S, et al. Validation of PHQ-2 and PHQ-9 to screen for major depression in the primary care population. Ann Fam Med. 2010;8(4):348–353. doi:10.1370/afm.1139

14. García-Campayo J, Zamorano E, Ruiz MA, Pérez-Páramo M, López-Gómez V, Rejas J. The assessment of generalized anxiety disorder: psychometric validation of the Spanish version of the self-administered GAD-2 scale in daily medical practice. Health Qual Life Outcomes. 2012;10(1):114.

15. USDA. U.S. Household Food Security Survey Module: Six-Item Short Form Economic Research Service. Published online September 2012. https://www.ers.usda.gov/media/8282/short2012.pdf

16. Zou G. A Modified Poisson Regression Approach to Prospective Studies with Binary Data. Am J Epidemiol. 2004;159(7):702–706. doi:10.1093/aje/kwh090

17. Coleman-Jensen A, Rabbitt M, Singh A. Household Food Security in the United States in 2019.; 2020.

18. Wolfson JA, Leung CW. Food Insecurity and COVID-19: Disparities in Early Effects for US Adults. Nutrients. 2020;12(6):1648.

19. Fitzpatrick KM, Harris C, Drawve G. Assessing Food Insecurity in the United States During COVID-19 Pandemic. Montana. 2020;34:3.

20. ERA Economics LLC. Economic Impacts of the COVID-19 Pandemic on California Agriculture. ERA Economics LLC; 2020. https://www.cfbf.com/wp-content/uploads/2020/06/COVID19_AgImpacts.pdf

21. Preliminary Data Brief. COVID-19 Farmworkers Study; 2020. http://covid19farmworkerstudy.org/survey/wp-content/uploads/2020/08/EN-COFS-Preliminary-Data-Brief_FINAL.pdf

22. Preliminary Data Brief: Oregon Fires Exacerbate COVID-19 Impact on Farmworkers’ Health, Housing, and Livelihoods. COVID-19 Farmworkers Study; 2020. http://covid19farmworkerstudy.org/survey/wp-content/uploads/2020/09/Preliminary-Data-Brief_OR_COFS_-9.22.20.pdf

23. Quandt SA, LaMonto NJ, Mora DC, Talton JW, Laurienti PJ, Arcury TA. COVID-19 Pandemic Among Immigrant Latinx Farmworker and Non-farmworker Families: A Rural-Urban Comparison of Economic, Educational, Healthcare, and Immigration Concerns. MedRxiv Prepr Serv Health Sci. Published online November 3, 2020:2020.10.30.20223156. doi:10.1101/2020.10.30.20223156

24. Wenham C, Smith J, Davies SE, et al. Women Are Most Affected by Pandemics—Lessons from Past Outbreaks. Nature Publishing Group; 2020.

25. Bandiera O, Buehren N, Goldstein MP, Rasul I, Smurra A. The Economic Lives of Young Women in the Time of Ebola: Lessons from an Empowerment Program. The World Bank; 2019.

26. Gausman J, Langer A. Sex and gender disparities in the COVID-19 pandemic. J Womens Health. 2020;29(4):465–466.

27. Wurth M, Bieber J, Klasing AM. Neglected and Unprotected: The Impact of the Zika Outbreak on Women and Girls in Northeastern Brazil. Human Rights Watch; 2017.

28. Albert PR. Why is depression more prevalent in women? J Psychiatry Neurosci JPN. 2015;40(4):219–221. doi:10.1503/jpn.150205

29. Salzwedel M, Liebman A, Kruse K, Lee B. The COVID-19 Impact on Childcare in Agricultural Populations. J Agromedicine. Published online 2020:1–5.

30. Hernandez T, Gabbard S. Findings from the National Agricultural Workers Survey (NAWS) 2015–2016. A Demographic and Employment Profile of United States Farmworkers. Dep Labor Employ Train Adm Wash Dist Columbia. Published online 2019.

31. Eskenazi B, Fahey CA, Kogut K, et al. Association of perceived immigration policy vulnerability with mental and physical health among US-born Latino adolescents in California. JAMA Pediatr. 2019;173(8):744–753.

32. Rivas R. California COVID-19 Farmworker Relief Package.; 2020. https://a30.asmdc.org/farmworker-relief-package

33. U.S. Department of Labor. Families First Coronavirus Response Act: Employee Paid Leave Rights.; 2020. https://www.dol.gov/agencies/whd/pandemic/ffcra-employee-paid-leave

34. Funk C, Tyson A. Intent to Get a COVID-19 Vaccine Rises to 60% as Confidence in Research and Development Process Increases. Pew Research Center; 2020. https://www.pewresearch.org/science/2020/12/03/intent-to-get-a-covid-19-vaccine-rises-to-60-as-confidence-in-research-and-development-process-increases/

35. SAGE Working Group. Report of the SAGE Working Group on Vaccine Hesitancy. World Health Organization; 2014. https://www.who.int/immunization/sage/meetings/2014/october/1_Report_WORKING_GROUP_vaccine_hesitancy_final.pdf

