## Supplemental Table 1 for "Impact of the COVID-19 Pandemic and Vaccine Hesitancy among Farmworkers from Monterey County, California"

| **Table S1.** Demographic and household characteristics, by sex and recruitment site, Monterey County COVID-19 Farmworker Study, July to November 2020, N=1115. | | | | | |
| --- | --- | --- | --- | --- | --- |
|  | Sex | |  | Recruitment site | |
|  | Male  N (%) or Mean ± SD | Female  N (%) or Mean ± SD |  | Clinic  N (%) or Mean ± SD | Outreach  N (%) or Mean ± SD |
| Total | 529 (47.4) | 586 (52.6) |  | 565 (50.7) | 550 (49.3) |
| Field workers |  |  |  |  |  |
| No | 132 (25.2) | 143 (24.6) |  | 139 (25.1) | 136 (24.7) |
| Yes | 391 (74.8) | 439 (75.4) |  | 416 (75.0) | 414 (75.3) |
| Age (in years) | 39.3 ± 13.5 | 40.0 ± 1.7 |  | 39.6 ± 12.5 | 39.7 ± 12.6 |
| 18-29 | 146 (27.6) | 131 (22.4)^a^ |  | 140 (24.8) | 137 (24.9) |
| 30-39 | 139 (26.3) | 135 (23.0) |  | 136 (24.1) | 138 (25.1) |
| 40-49 | 117 (22.1) | 181 (30.9) |  | 163 (28.9) | 135 (24.6) |
| 50+ | 127 (24.0) | 139 (23.7) |  | 126 (22.3) | 140 (25.5) |
| Education |  |  |  |  |  |
| Primary school or less | 212 (40.1) | 280 (47.9)^a^ |  | 277 (49.1) | 215 (39.1)^b^ |
| More than primary school | 317 (59.9) | 305 (52.1) |  | 287 (50.9) | 335 (60.9) |
| Language |  |  |  |  |  |
| Spanish | 440 (83.2) | 508 (86.7) |  | 440 (81.4) | 488 (88.7)^b^ |
| English | 34 (6.4) | 23 (3.9) |  | 21 (3.7) | 36 (6.6) |
| Indigenous | 55 (10.4) | 55 (9.4) |  | 84 (14.9) | 26 (4.7) |
| Birthplace |  |  |  |  |  |
| Mexico | 434 (82.0) | 495 (84.5) |  | 486 (86.0) | 443 (80.6)^b^ |
| United States | 78 (14.7) | 64 (10.9) |  | 49 (8.7) | 93 (16.9) |
| Other | 17 (3.2) | 27 (4.6) |  | 30 (5.3) | 14 (2.6) |
| Years in US | 20.7 ± 12.7 | 20.6 ± 9.8 |  | 19.9 ± 11.2 | 21.5 ± 11.2^b,c^ |
| <20 years | 215 (47.7) | 241 (46.3) |  | 267 (51.8) | 189 (41.4) |
| ≥20 years | 236 (52.3) | 280 (53.7) |  | 248 (48.2) | 268 (58.6) |
| Marital status |  |  |  |  |  |
| Not married or living as married | 176 (33.3) | 235 (40.1)^a^ |  | 225 (39.8) | 186 (33.9)^b^ |
| Married or living as married | 352 (66.7) | 351 (59.9) |  | 340 (60.2) | 363 (66.1) |
| Residence |  |  |  |  |  |
| Salinas | 196 (37.1) | 296 (50.5)^a^ |  | 263 (46.6) | 229 (41.6)^b^ |
| Greenfield | 165 (31.2) | 151 (25.8) |  | 217 (38.4) | 99 (18.0) |
| Other | 168 (31.8) | 139 (23.7) |  | 85 (15.0) | 222 (40.4) |
| Annual household income |  |  |  |  |  |
| <$25,000 | 221 (44.1) | 339 (60.8)^a^ |  | 291 (54.3) | 269 (51.4) |
| ≥$25,000 | 280 (55.9) | 219 (39.3) |  | 245 (45.7) | 254 (48.6) |
| Household crowding |  |  |  |  |  |
| ≤2 persons per bedroom | 325 (61.4) | 383 (65.4) |  | 337 (59.7) | 371 (67.5)^b^ |
| >2 persons per bedroom | 204 (38.6) | 203 (34.6) |  | 228 (40.4) | 179 (32.6) |
| Live with unrelated roommates |  |  |  |  |  |
| No | 395 (74.7) | 514 (87.7)^a^ |  | 450 (79.7) | 459 (83.5) |
| Yes | 134 (25.3) | 72 (12.3) |  | 115 (20.4) | 91 (16.6) |
| Children under 18 living in home |  |  |  |  |  |
| No | 172 (32.6) | 106 (18.1)^a^ |  | 125 (22.1) | 153 (27.9)^b^ |
| Yes | 356 (67.4) | 480 (81.9) |  | 440 (77.9) | 396 (72.1) |
| ^a^Difference between male and female participants (p<0.05).  ^b^Difference between clinic-recruited and outreach-recruited participants (p<0.05).  ^c^Among those not born in the United States (n=973) | | | | | |
